# A new form of hereditary iron overload unlinked to known hereditary haemochromatosis genes

**DOI:** 10.1101/2024.05.10.24307148

**Authors:** Daniel F. Wallace, V. Nathan Subramaniam

**Author notes:** Corresponding Authors:Daniel Wallace, V. Nathan Subramaniam.

## Abstract

Hereditary iron overload conditions are normally caused by mutations in the *HFE* gene or other genes involved in the regulation of iron homeostasis. We describe a family with an iron overload condition with apparent autosomal dominant inheritance and phenotypic similarities to the classical form of ferroportin disease. The condition is characterised by elevated serum ferritin levels with normal or mildly elevated transferrin saturation and prominent iron deposition in both Kupffer cells and hepatocytes. Serum hepcidin levels were elevated in affected members of the pedigree and correlated with serum ferritin concentration. No pathogenic variants were identified in *HFE* or *SLC40A1*, the gene encoding ferroportin. Whole genome linkage analysis showed maximal linkage of the iron overload phenotype to regions on chromosomes 1, 3, 12, 18 and 19, indicating that affected members of this family do not have a variant form of ferroportin disease and are likely to have a new form of hereditary iron overload due to defects in another gene.

## Introduction

Hereditary haemochromatosis (HH) is normally caused by homozygosity for pathogenic mutations in the *HFE* (homeostatic iron regulator) gene (1). Other autosomal recessive forms of HH can be caused by mutations in other genes involved in the regulation of iron homeostasis including hemojuvelin (*HJV*), hepcidin (*HAMP*) and transferrin receptor 2 (*TFR2*) (2). Autosomal dominant forms of iron overload have been associated with mutations in the cellular iron exporter ferroportin (encoded by *SLC40A1*) (3), with a single case linked to a mutation in the iron responsive element (IRE) located in the 5’UTR of the H-ferritin gene (*FTH1*) (4).

Heterozygosity for pathogenic variants in *SLC40A1* can lead to one of two phenotypes. Loss of function variants, that cause defects in the iron transport activity of ferroportin lead to “classical ferroportin disease”, a form of iron overload characterised by high serum ferritin, normal or mildly elevated transferrin saturation and iron accumulation predominantly in Kupffer cells in the liver (5). Gain of function variants, that cause disruption to hepcidin-mediated ferroportin down-regulation lead to a phenotype similar to HFE-related haemochromatosis, characterised by elevated serum ferritin and transferrin saturation, with iron accumulation predominantly in hepatocytes (5).

We describe a multi-generation family with an iron overload condition that has an apparent autosomal dominant inheritance pattern. The characteristics of the condition are similar to the classical form of ferroportin disease, with elevated serum ferritin levels, normal or mildly elevated transferrin saturation and prominent iron deposition in both Kupffer cells and hepatocytes. We have investigated the genetic basis of iron overload in this family and conclude that the genetic basis of the condition is unlinked to currently known haemochromatosis genes.

## Materials and Methods

### Patients

This study was approved by and performed in accordance with the ethical standards of the QIMR Berghofer Medical Research Institute Human Research Ethics Committee, the Queensland University of Technology Human Research Ethics Committee and with the Helsinki Declaration of 1975, as revised in 1983. Informed and written consent was obtained from the participants for all studies described in this report.

An Australian family with multiple members affected with iron overload was identified. Blood samples were taken, serum iron parameters, including serum iron, transferrin saturation and serum ferritin were measured and *HFE* genotypes for the p.C282Y, p.H63D and p.S65C variants were determined. Blood samples were processed to separate plasma and buffy coats. Liver biopsy samples were obtained from the proband and his affected nephew and stained with Perls’ Prussian blue for iron.

### Genetic studies

Genomic DNA was extracted from buffy coats and Sanger DNA sequencing of the coding sequence and intron/exon boundaries of the *SLC40A1* gene and the IREs of the *FTL* and *FTH1* genes was performed in the proband as described previously (6). Whole genome single nucleotide polymorphism (SNP) genotyping was done in 15 members of the pedigree using Illumina Human610-Quad BeadChips. Linkage analysis was performed using Superlink-Online SNP 1.1.

### Hepcidin measurements

Serum hepcidin concentrations were measured in plasma samples using a liquid chromatography coupled with tandem mass spectrometry (LC-MS/MS) method (7) and correlated with serum ferritin concentration measurements.

## Results and Discussion

The proband from a large Australian family with southern European ancestry presented with an elevated serum ferritin concentration of 1907 µg/L and mildly elevated transferrin saturation of 50.4%. *HFE* gene testing revealed normal genotypes for the p.C282Y, p.H63D and p.S65C variants. A liver biopsy was done, and Perls’ Prussian blue staining revealed grade II iron accumulation in hepatocytes with prominent iron deposition in Kupffer cells. Other family members, including two siblings and a nephew of the proband were identified with elevated serum ferritin levels (1212 µg/L, 828 µg/L and 1163 µg/L, respectively), and iron overload was confirmed in the nephew by Perls’ Prussian blue staining of liver biopsy sections, revealing a similar pattern of iron deposition in hepatocytes and Kupffer cells (Figure 1). Several other members of the pedigree were identified with mild to moderate elevations in serum ferritin across three generations. Full pedigree data is available upon request from the corresponding authors. Some members of the pedigree were heterozygous for the *HFE* p.H63D variant, but none had *HFE* genotypes that are associated with HH. Sequencing of the entire coding region and splice sites of the *SLC40A1* gene and the IREs of *FTH1* and *FTL* revealed no pathogenic variants in the proband, suggesting that the iron overload and hyperferritinaemia present were unrelated to variants in these genes. Serum hepcidin concentration was elevated in the proband and there was a positive correlation with serum ferritin among all pedigree members (Figure 2), suggesting that the iron overload phenotype is likely unrelated to defects in hepcidin regulatory pathways. The number of individuals with hyperferritinaemia, and its occurrence in potentially three generations of the pedirgree, combined with the pattern of liver iron loading observed in two pedigree members led us to hypothesise that the iron overload condition had autosomal dominant inheritance and was phenotypically similar to classical ferroportin disease but unrelated to variants in the *SLC40A1* gene.

**Figure 1.**
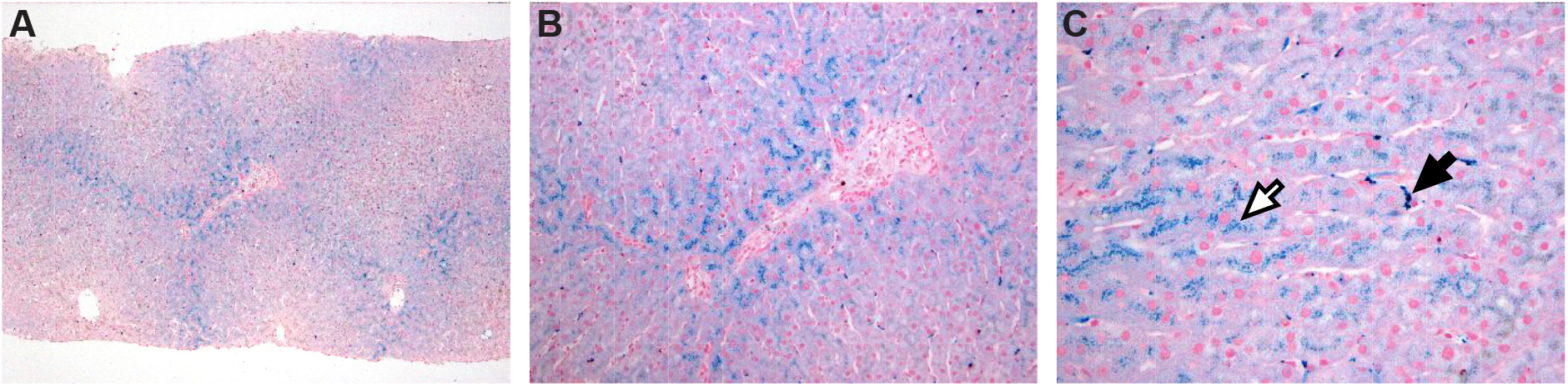
Liver biopsy sections showing prominent iron deposition in hepatocytes and Kupffer cells. (A) Low (40x), (B) medium (100x) and (C) high (400x) magnification images of liver biopsy sections from the proband’s nephew stained with Perls’ Prussian blue. Periportal iron loading can be seen with prominent iron deposits in both hepatocytes (white arrow) and Kupffer cells (black arrow).

**Figure 2.**
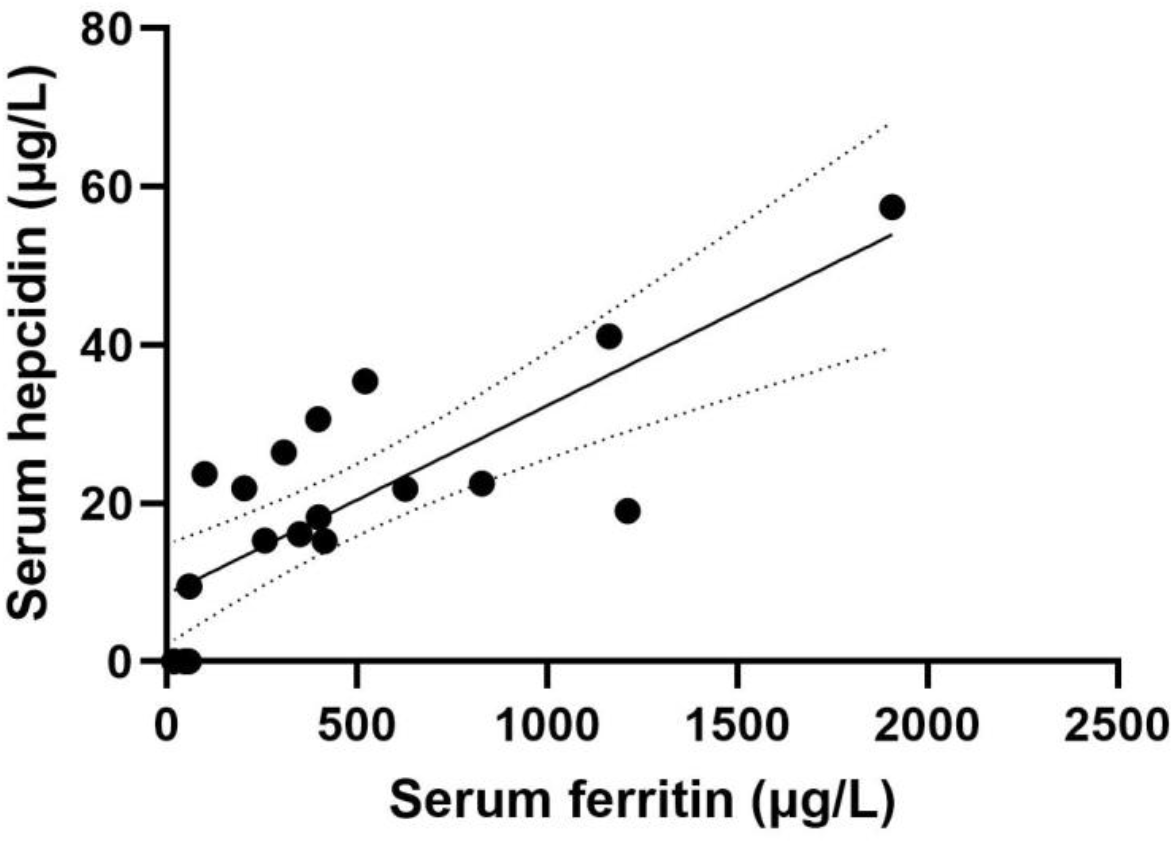
Serum hepcidin correlates with serum ferritin in pedigree members. Serum ferritin was plotted against serum hepcidin measured in the plasma of 19 pedigree members, showing a highly significant positive correlation (r=0.79, p <0.0001). Dashed lines represent 95% confidence intervals.

In an attempt to identify a genetic cause for iron overload, 15 members of the pedigree were genotyped using whole genome SNP microarrays. Linkage analysis was performed, assuming an autosomal dominant inheritance pattern, including the four most clearly affected and the three most clearly unaffected members of the pedigree. The results of the linkage analysis showed maximal linkage of the iron overload phenotype to regions on chromosomes 1, 3, 12, 18 and 19 (Figure 3), with haplotypes in these regions that are shared among the four most clearly affected members of the pedigree but absent in the three most clearly unaffected members. These regions encompass a total of 93 megabases and contain over 1500 canonical genes. Importantly, these genomic intervals do not contain the *SLC40A1* gene indicating that affected members of this pedigree do not have a variant form of ferroportin disease. We conclude that this family has an iron overload condition with probable autosomal dominant inheritance that is unlinked to the currently known haemochromatosis genes. Investigations are underway to identify the molecular basis for iron overload in this family.

**Figure 3.**
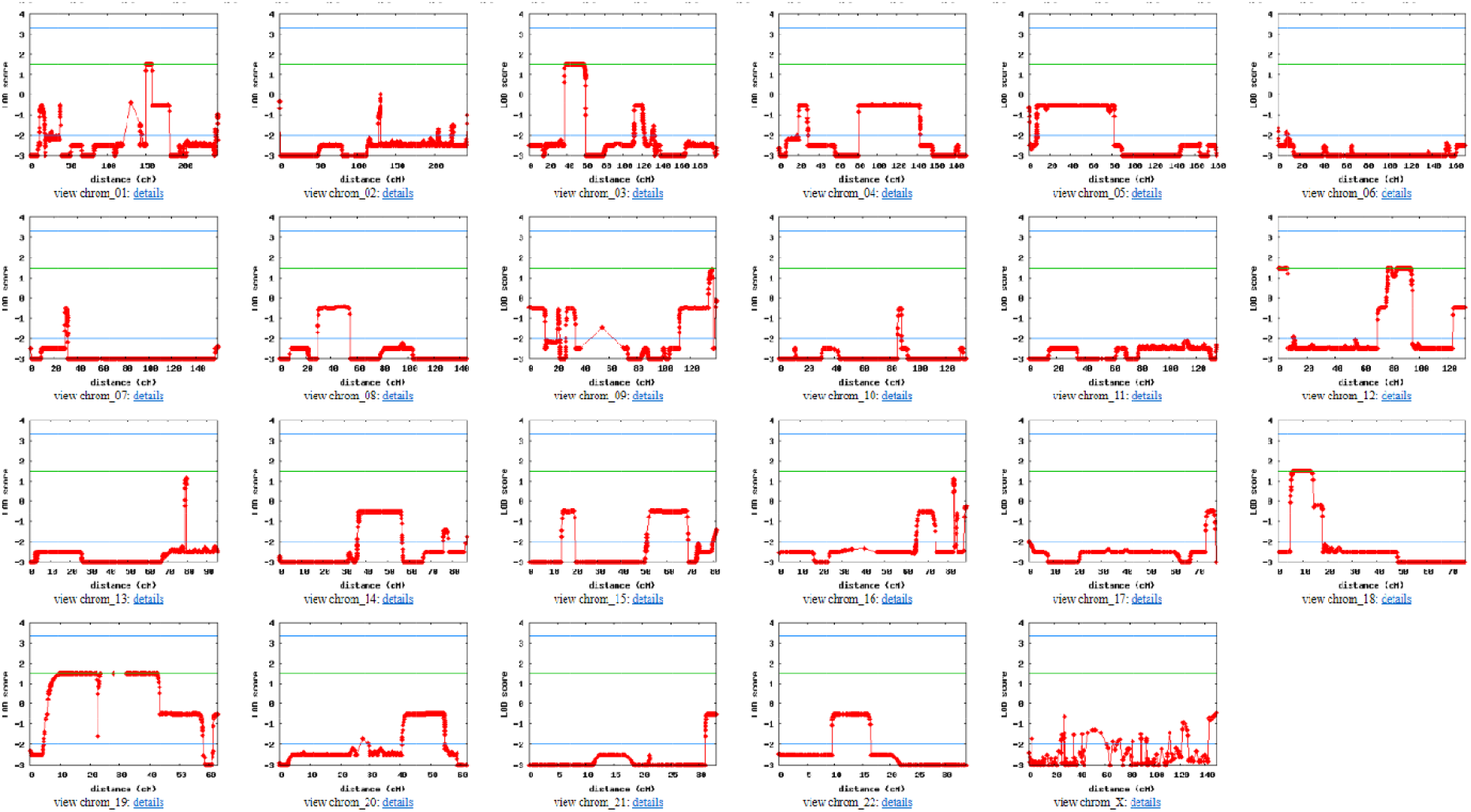
Linkage analysis revealed maximal linkage of iron overload condition to regions of chromosomes 1, 3, 12, 18 and 19. Linkage analysis was performed, assuming autosomal dominant inheritance, using whole genome SNP data from the four most clearly affected members and the three most clearly unaffected members of the pedigree using the Superlink-Online SNP 1.1 program. The green line represents maximal linkage in this analysis. Maximal linkage was observed in regions of chromosomes 1, 3, 12, 18 and 19.

## Data Availability

All data produced in the present study are available upon reasonable request to the authors

